# To resect or not to resect? Unbiased performances of single and combined biomarkers in intra-operative corticography for tailoring during epilepsy surgery

**DOI:** 10.1101/2019.12.26.19015883

**Authors:** Matteo Demuru, Stiliyan Kalitzin, Willemiek Zweiphenning, Dorien van Blooijs, Maryse van ’t Klooster, Pieter Van Eijsden, Frans Leijten, Maeike Zijlmans, on behalf of the RESPECT group

## Abstract

**Objective:** Signal analysis biomarkers, in an intra-operative setting, may be complementary tools to guide and tailor the resection in drug-resistant epilepsy patients. Unbiased assessment of biomarker performances are needed to evaluate their clinical usefulness and translation. We defined a realistic ground-truth scenario and compared the effectiveness of different biomarkers alone and combined to localize epileptogenic tissue.

**Methods:** We investigated the performances of univariate, bivariate and multivariate signal biomarkers applied to 1 minute inter-ictal intra-operative electrocorticography to discriminate between electrodes covering normal or pathologic activity in 47 drug-resistant people with epilepsy (temporal and extra-temporal) who had been seizure-free one year after the operation.

**Results:** The best result using a single biomarker was obtained using the phase-amplitude coupling measure for which the epileptogenic tissue was localized in 16 out of 47 patients. Combining the whole set of biomarkers provided an improvement of the performances: 20 out of 47 patients. Repeating the analysis only on the temporal-lobe resections we reached a sensitivity of 93% (28 out of 30) combining all the biomarkers.

**Conclusion:** We suggest that the assessment of biomarker performances on a ground-truth scenario is required to have a proper estimate on how biomarkers translate into clinical use. Phase-amplitude coupling seems the best performing single biomarker and combining biomarkers improves localization of epileptogenic tissue. However, sensitivity achieved is not adequate for the usage as a tool in the operation theater, but it can improve the understanding of pathophysiological process.

## Introduction

Epilepsy is a disorder which affects the life of around 50 millions of people worldwide^1^. One third of epileptic patients are drug-resistant^2,3^. Epilepsy surgery provides a potential cure for these patients. The success of surgery is linked to the mapping of the epileptogenic zone (EZ), the minimum cortical area that needs to be resected to achieve seizure freedom^4^.

Intra-operative electro-corticography (ioECoG) can be used during surgery to assess and adjust the boundaries of the proposed resection area. Typically, tissue showing inter-ictal epiletiform activity (i.e. spikes) is the main factor to guide the definition of the boundaries^5^. The complete removal of spikes has been correlated with a good post-surgical seizure outcome^6–11^, but good outcomes are also seen in cases where not all spikes are removed, and bad outcomes in cases where no spikes were seen^12–14^. Ictiform spike patterns are a more specific marker than sporadic spikes^11^. Recently, it was found that high frequency oscillations (HFOs; above 80 Hz) may be more specific predictors of outcome, especially when still present after the resection^15–17^.

Inter-ictal recordings are richer in content than only spikes, that is why more sophisticated approaches exploiting different features of the recorded signals were suggested as complementary methods to spikes^18–20^. During the past years a plethora of biomarkers based on signal analysis techniques has been developed. These biomarkers can be conceptually subdivided in three categories: univariate, bivariate and multivariate methods. Univariate methods provide information related to each signal separately, bivariate methods investigate the relationship, most typically correlation, between each couple of signals (known as functional connectivity^21^), while multivariate methods estimate global relationship between all the signals available. There are other distinctions to differentiate the methods for each of the categories: for example when calculating signal correlations whether the calculation of the biomarker is amplitude or phase-based, time or frequency based, undirected or directed, linear or nonlinear^18–20^.

Several studies have shown that using certain biomarkers the epileptogenic tissue can be distinguished from the healthy tissue based on inter-ictal intracranial recordings^22–29^, but only statistically on the aggregate group level. A biomarker set is needed that discriminates well between tissue that needs to be surgically removed and tissue that should not at the patient level, which can thus be applied for clinical care.

However, clinical translation of biomarkers is hampered by high inter-subject variability, which hinders their generalizability. This problem is compounded by the lack of an objective (and commonly accepted) way to assess the biomarker performances in comparison to a ground truth.

The definition of a ground truth scenario is a delicate issue that is most of the time overlooked. Typically, a tissue (or recording sites) is labeled into one of the two classes normal/pathological based on pre-resection recordings; where a tissue is defined as ‘normal’ or epileptogenic depending on some criteria, like inclusion or not in the seizure onset zone (SOZ), inclusion or not in the resected area, that may or may not correspond well to the actual epileptogenic zone. For instance, resections may span over normal tissue for surgical constraints rather than suspicious epileptogenic activity.

Furthermore, the incomplete coverage of the whole brain for the recordings may bias the definition of the two classes. For example, the absence of any detection of pathological channels may be due to the poor sensitivity of the biomarker but also to a misplaced recording arrangement (i.e. we are not recording in the proper spot). Therefore, computing sensitivity and specificity in this way does not provide a reliable assessment of the biomarker performances.

We aimed to develop a pragmatic approach to overcome these issues and work on a realistic ground truth scenario. This is possible thanks to the nature of our data: we have both pre-resection and post-resection recordings, information related to the resected area and good seizure outcome patients. We reasoned that we can claim that EZ was sufficiently removed in those patients who become seizure-free after the resection, such that we may use the post-resection recordings to compute a reliable biomarker reference value for normal tissue (i.e. not able to trigger a seizure). Comparing this reference to the pre-resection biomarker values, computed on the resection area, makes it possible to have a proper estimate of the performances of the biomarker.

We would like to point out that using this analysis approach the only sound estimate to assess the biomarker performances is the sensitivity (i.e. the number of subjects who have at least a biomarker value computed in the resected areas during the pre-resection recordings higher than the reference biomarker value), while specificity (i.e. the number of subjects who have all biomarker value in the not-resected area during the pre-resection recordings lower or equal the reference biomarker) is biased since we cannot have full coverage of the brain recording not-resected areas.

We investigated, in our ground-truth scenario, the performances of different biomarkers that were already used in previous studies^23–25,27–35^. We defined our biomarker pool with the attempt to be exhaustive according to the three different categories univariate, bivariate and multivariate biomarkers. We combined all the biomarkers together, given that they potentially carry different information, and assessed the performance of our multi-feature biomarker.

## Methods

### Patients

We selected patients from a retrospective database of refractory epilepsy patients (RESPect) who underwent ioECoG-tailored resective surgery at the University Medical Center in Utrecht, the Netherlands, between 2008 and 2018. The RESPect database contains both retrospective data and prospectively gathered data. Informed consent is being obtained for prospective part. For the retrospective part, the Medical Ethical committee of the UMC Utrecht agreed that informed consent would not be requested. We consecutively anonymized, visually assessed and annotated (i.e. bad channels marked, artefact marked, good segments of the signal marked) and imported into the brain imaging data structure (BIDS)^36^ (see https://github.com/suforraxi/ieeg_respect_bids). We included patients if (1) the data was anonymized, visually assessed, annotated and imported in BIDS (2) at least one minute artefact free pre- and post-resection ECoG recordings was present (3) post-surgical seizure outcome after 1 year was available, (4) the recording grids format was 4 x 5 with or without additional strips, (5) pictures pre- and post-resection were present to label the electrodes (resected or not resected), (6) the ECoG was recorded at a 2,048 Hz sampling frequency, (7) patients were not included in the HFO trial^37^, (8) one year good seizure outcome (Engel 1A or 1B) after the operation.

These criteria restricted our dataset to 47 patients: 30 of those are temporal lobe who had hippocampectomy and the remaining 17 were extra-temporal patients. We defined these 47 patients as improved. We defined a subset of patients as ‘cured’ if after one year they belonged to Engel 1A class and they stopped the medication.

### Data acquisition

IoECoG signals were recorded for clinical purposes using 4 x 5 electrode grids and 1 x 6 or 1 x 8 electrode strips (Ad-Tech, Racine, WI) placed directly on the cortex. The grids and strips consist of platinum electrodes with 4.2 mm^2^ contact surface, embedded in silicone, and 1 cm inter-electrode distances. Recordings were made with a 64-channel EEG system (MicroMed, Veneto, Italy) at 2,048 Hz sampling rate using an anti-aliasing filter at 538 Hz. The signal was referenced to an external electrode placed on the mastoid. Grids and electrode strips were placed in multiple arrangements before and after resection. Propofol was used to induce general anesthesia and maintained using a propofol infusion pump. Propofol was interrupted until a continuous ECoG background pattern was achieved.

### Data selection

For each recording arrangement (from here on ‘situation’) we visually selected one minute artefact free recording starting at the end of the recording and going backwards. This was done to minimize the propofol effect. We considered all the situations before the first resection was performed and all the post-resection situations (i.e. the resection was completely finished).

### Data preprocessing and processing

The recordings from channels with visually marked noise (double checked by at least two people in a common reference montage) were excluded. The data was then re-referenced using a bipolar montage. The bipolar montage for the grid was computed both along the horizontal and vertical directions of the grid. This was done in order to take into account possible different orientations of the sources underneath and to optimally use all electrodes. For each situation the selected minute was divided in 5 second segments and the following preprocessing steps were applied independently for each segment: detrending, demeaning and z-score transformation. Depending on the specific measure, additional pre-processing steps were applied (see <u>Biomarkers</u> section for details). If the specific measure required filtering a finite impulse response (FIR) filter was used. For every univariate measure (Auto-Regressive Residual Modulation, and phase-amplitude coupling, see <u>Biomarkers</u>) we averaged the values across the segments to obtain a unique value for each bipolar channel across the situation. For bivariate and multivariate measures, producing functional connectivity matrix in each time segment, we first averaged in order to have one value per bipolar channel; second we averaged these values across time segments. Granger Causality (GC) was a special case because the multivariate model was fitted pooling the segments all together which resulted in one functional connectivity matrix (i.e. no need to average across the segments).

If the functional connectivity matrix was obtained from a directional measure (non linear correlation coefficient h^2^,GC, short-time direct Directed Transfer Function, sdDTF see <u>Biomarkers</u>) we considered the out-strength (i.e. the effect that a channel has on the other channels).

### Identification of resected tissue

Electrodes were classified into resected or non-resected using photographs taken during surgery. Furthermore, because we applied a bipolar montage we labeled a bipolar channel as resected if both monopolar channels were included in the resection area; we labeled a bipolar channel not-resected if both the monopolar channels were excluded from the resected area and we did not consider intermediate cases (bipolar derivations for which one monopolar channel was resected and the other not).

### Measuring effect across all the channels

We considered for each biomarker all values computed in the pre-resection situations for channels that were eventually resected (from now on ‘pre-resection resected channels’), and we compared them with the channel values computed on all post-resection situations in cured patients to assess if the different biomarkers could detect an effect on a group level. We tested for differences in the distributions with a two-sample one-sided Kolmogorov-Smirnov test (testing pre-resection resected values > post-resection values).

### Measuring effect using maximum per patient

All biomarkers that showed a significant difference (p < 0.01, no correcting for multiple comparison) in the previous analysis were further analyzed. We calculated the maximum value of the biomarker across all pre-resection situations in resected channels of improved patients and the maximum value across all post-resection situations in cured patients. Then, we compared the distributions of maxima across patients between pre-resection resected channels and post-resection channels using a two-sample one-sided Kolmogorov-Smirnov test (testing pre-resection resected values > post-resection values).

We defined a threshold to discriminate between pathological and healthy tissue by taking the maximum value of the biomarker across all channels of all post-resection situations in all cured patients. We reasoned that if a patient becomes seizure-free without medication after surgery, it means that the operation was successful: enough tissue was removed and the remaining tissue can be considered not able to generate seizures. Therefore, measuring the biomarker in this tissue (what is left after resection, post-resection situations) can give an estimate of ‘normal’ values of the biomarker. Choosing a threshold as the maximum across channels/situations/patients represents a way to define a ‘universal’ threshold that can be applied to discriminate between normal and epileptogenic tissue even for new patients. We measured the performances of each biomarker with the sensitivity by counting how many patients had at least one value of the pre-resection resected values above the threshold.

Furthermore, we defined a ‘cumulative’ biomarker combining together all the biomarkers. Specifically, we considered a true positive if any of the biomarkers is above the respective threshold. Finally, we repeated the analysis (recomputing the thresholds) considering the subgroups of temporal and extra-temporal patients.

### Mesiotemporal versus neocortical channels

For temporal patients, the first three electrodes of the electrode strip directed at the mesiotemporal structure (hippocampus, amygdala and enthorhinal cortex) were classified as mesiotemporal channels and the other channels (i.e. grid) were classified as neocortical channels. We compared the values of hippocampal channels and neocortical channels using a two-sample one-sided Kolmogorov-Smirnov test (testing mesiotemporal channel values > neocortical channel values) in order to understand the effect of the anatomical structure on the overall result.

### Univariate Biomarkers

#### Auto-Regressive Residual Modulation

The auto-regressive residual modulation (ARRm) provides the amount of non-harmonicity in the signal quantified as the high residual variation after auto-regressive modelling^30,31^. It has been shown that brain tissue with high non-harmonicity corresponds to areas with high frequency oscillations (HFOs) which in turn may be an indication of epileptogenic tissue^38–42^. Following Geertsema’s work^31^ we defined the ARRm parameters as : (1) window length of 40 samples, which with a sample frequency of 2048 Hz, corresponds to approximately 20 ms; (2) consecutive 50% overlapping windows. For the detailed formula see Appendix.

#### Phase Amplitude Coupling

Phase-amplitude coupling (PAC) is a form of cross-frequency coupling^43^ where the amplitude of a higher frequency oscillation is modulated by the phase of a lower frequency oscillations. Recent studies^32–35^ have shown that high PAC values are related with the SOZ. There are many proposed methods to estimate PAC^44–50^ and different parameter choices that can be made (i.e. the low and high frequency interval where estimate the phase and amplitude). There is no systematic study evaluating the performances of PAC in localizing the epileptogenic tissue using either different PAC implementations or the different parameters choices. We decided to investigate our dataset computing PAC between the modulating phase of theta band activity (4-8 Hz) and the amplitude of gamma activity (30-80 Hz) because this frequency band pairs were successfully investigated in recent epilepsy related studies^32–35^. For the detailed formula see Appendix.

### Bivariate biomarkers

#### Phase Locking Value

Phase locking values^51^ (PLV) is a non linear bivariate measure quantifying frequency-specific phase synchronization between two signals. Mormann and colleagues^27^ have been one of the first group to show how mean phase coherence (another name for PLV) can correctly lateralize the side of the epileptic focus using inter-ictal ECoG recordings. We computed PLV in the gamma frequency band (30-80Hz), see the formula in the Appendix.

#### Phase Lag Index

The phase lag index^52^ (PLI) is a bivariate measure quantifying the asymmetry of the distribution of the phase differences between two signals. Van Dellen et al.^24^ investigated inter-ictal ECoG using PLI in temporal lobe patients. They showed that PLI was related to disease history. Moreover, van Diessen et al.^25^ found that network based PLI quantities (strength and eigenvector centrality) in theta and gamma frequency bands, were associated with areas with HFOs and SOZ in temporal lobe patients. We chose to compute PLI in gamma band (30-80Hz) using the formula in the Appendix.

#### Non linear correlation coefficient

The non linear correlation coefficient 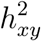 between signal *x* and *y* is an extension of the linear correlation coefficient that captures both linear and non linear interactions. It has been widely used to analyze brain signals in the field of epilepsy (see recent reviews^18,19^). Of note, the work of Bettus and colleagues^28^ showed that *h*^2^ provided information related to the localization of the epileptogenic focus using inter-ictal ECoG recordings. In this latter work the effect measured by *h*^2^ was significant for theta, alpha, beta and gamma bands and mostly independent to inter-ictal spiking. We computed *h*^2^ in gamma band (30-80Hz) using the formula in the Appendix. We looked also at the possible delayed effect computing shifting one signal compared to the other for different delays ([−0.0332s 0.0332s] in steps of 0.0083s) and we chose the delay which gave the maximum *h*^2^.

#### Granger Causality (time-domain)

Granger causality (GC) methods are statistical approaches based on auto-regressive modelling determined on the data that estimated the amount directed relationship among different time-series. In a bivariate scenario, one time-series *x* ‘Granger cause’ another time series *y* if the inclusion of past values of *x* reduces the variance of the modelling error compared to modelling error using only *y* past values. This can be generalized to a multivariate scenario where the reduction of modelling error of the multivariate model is used instead (see Blinowska^53^ for detailed review).

Park and colleagues^29^ successfully applied time based multivariate Granger causality to inter-ictal ECoG recordings showing that ictal networks can be inferred from inter-ictal recordings. They showed that there was a significant correlation between the epileptogenic location inferred using ictal recordings (i.e. defined by a neurologist team) with the location pointed out using GC on inter-ictal recordings. We applied the analysis pipeline suggested by Park et al.^29^, therefore we added the first order differentiation as an extra step in the pre-processing analysis before to compute the z-scores, furthermore the Akaike’s Information Criterion^54^ was used to select the optimal model order. See the Appendix for formulas.

#### short-time direct Directed Transfer Function

An extension of Granger causality methods to the frequency domain is the directed transfer function^55^. It estimates the causal (in Granger sense, reduction of the modelling error) influence a time-series *x* exerts on a *y* time-series in a multivariate modelling of the time-series. Short-time DTF (sdDTF) represents a further development of the DTF in order to capture the dynamic changes of the causal relationship^53^.

Zweiphenning and colleagues^23^ using sdDTF on ioECoG recordings observed that the out-strength (i.e. quantification of the ‘driving’ behaviour of a channel) of a channel in high-frequency bands (gamma and ripple band) matched the resected channels in patients with a seizure-free outcome. We computed sdDFT following the Zweiphenning’s pipeline, therefore we chose a model order of 30 samples, since this model order gave the best results. See Appendix for details.

### Code Implementation

All the code is available at https://github.com/suforraxi/multiple_biomarkers. We used MATLAB (Release R2019a, The MathWorks, Inc., Natick, Massachusetts, United States.) as a software framework plus the following toolboxes fieldtrip^56^, SIFT^57,58^ (for the computation of sdDTF) and MVGC^59^ (for the computation of GC).

## Results

### Patient description

Table 1 shows the patients characteristics. Our dataset consisted of 47 drugs-resistant epilepsy patients of whom 23 were male. Thirty of these patients were temporal patients who underwent hippocampectomy, while the remaining were extra-temporal. All the patients have a good seizure outcome after one year (Engel 1A). Thirteen patients could successfully withdraw all medication after surgery (cured patients) ; 16 patients managed to control seizures with a lower dosage of anti-epileptic medication and 18 kept the same dosage of medication. The primary pathology diagnosis is reported in Table 1. The majority of the patients (17) had a low grade tumor (WHO I + II) diagnosis and 7 patients had focal cortical dysplasia, 7 cavernoma and 7 gliosis/scar diagnosis classes. There were 5 patients with mesiotemporal sclerosis, 2 patients with cortical malformation development, one patient with tubero-sclerosis and one patient with no abnormalities. The total number of bipolar channels in the post-resection recordings were 1864, while 1138 channels were recorded during the pre-resection phase were eventually resected (labeled as resected channels). For our analysis we did not use the no-resected channels (1754) and the channels (417) for which we could not assign a label (cut channels) from the pre-resection recordings. On average we have around 40 channels per subject recorded in the post-resection and about 24 channels labeled as resected in the pre-resection recordings.

**Table 1.**
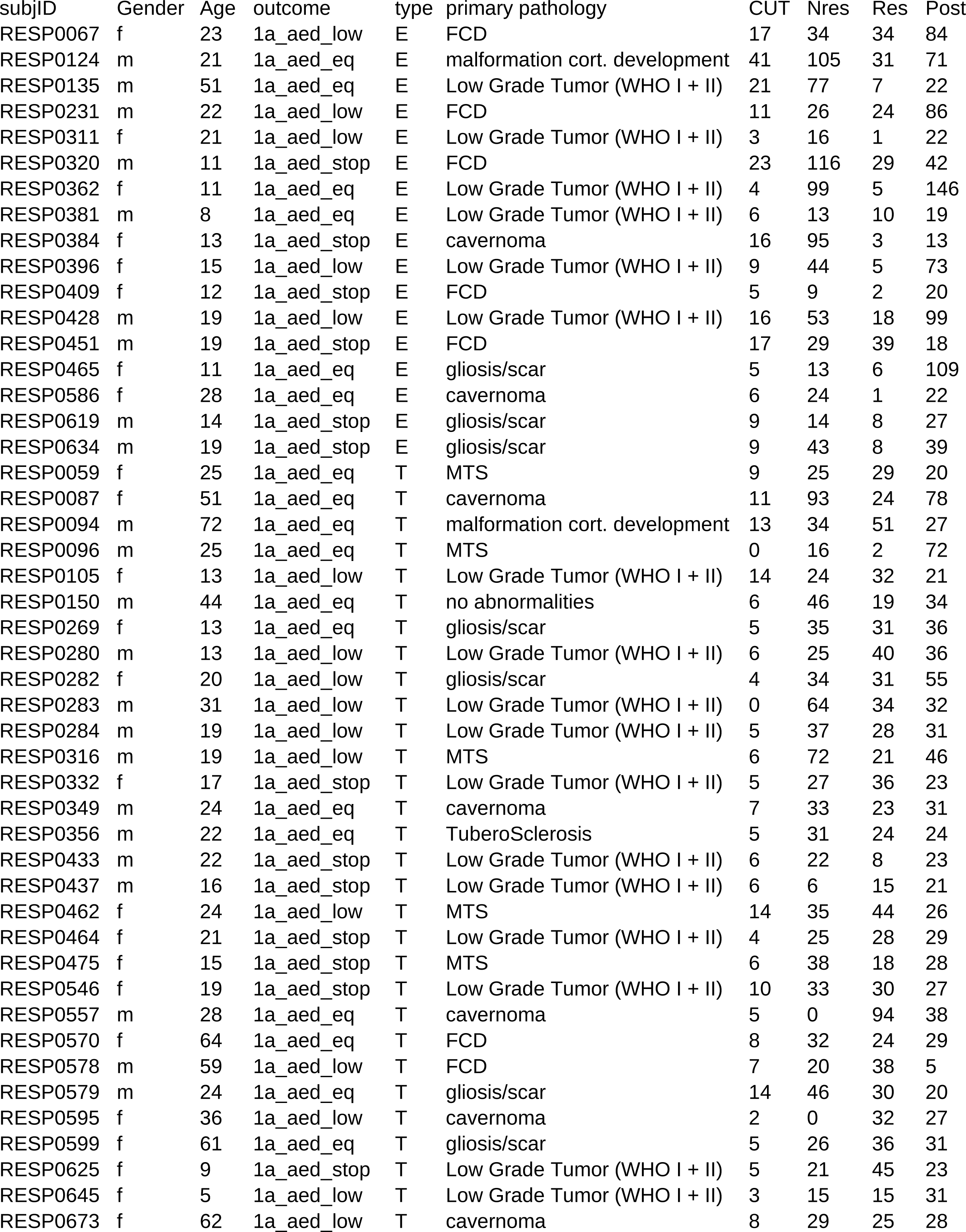
Patient characteristics. The complete dataset consists of 47 patients. The variable *subjID* is the code used to identify the subjects. The variable *outcome* represents the seizure outcome one year after surgery. This is identified by a code composed by the Engel class and the amount of medication after surgery (i.e. 1a_aed_stop means 1A Engel class who stop the medication after surgery, 1a_aed_low means that the medication was lowered and 1a_aed_eq means that the amount of medication was the same as before surgery). The variable *type* represents the type of epilepsy (E for Extra-Temporal, T for Temporal). The variable *primary pathology* represent the primary pathology and could be one of the following: low grade tumor (WHO I + II), mesiotemporal (MST), focal cortical dysplasia, carvernoma, gliosi/scar, malformation of cortical development, no abnormalities. The variable CUT represents the number of bipolar derivations where one electrode of the bipolar channel is resected and the other is not in the pre-resection situations. The variable Nres counts the number of not-resected channels in the pre-resection situations. The variable Res counts the number of resected channel in the pre-resection situations and finally the variable Post counts the number of bipolar channels in the post-resection situations.

characteristic\_tbl
| subjID | Gender | Age | outcome | type | primary pathology | CUT | Nres | Res | Post |
| --- | --- | --- | --- | --- | --- | --- | --- | --- | --- |
| RESP0067 | f | 23 | 1a_aed_low | E | FCD | 17 | 34 | 34 | 84 |
| RESP0124 | m | 21 | 1a_aed_eq | E | malformation cort. development | 41 | 105 | 31 | 71 |
| RESP0135 | m | 51 | 1a_aed_eq | E | Low Grade Tumor (WHO I + II) | 21 | 77 | 7 | 22 |
| RESP0231 | m | 22 | 1a_aed_low | E | FCD | 11 | 26 | 24 | 86 |
| RESP0311 | f | 21 | 1a_aed_low | E | Low Grade Tumor (WHO I + II) | 3 | 16 | 1 | 22 |
| RESP0320 | m | 11 | 1a_aed_stop | E | FCD | 23 | 116 | 29 | 42 |
| RESP0362 | f | 11 | 1a_aed_eq | E | Low Grade Tumor (WHO I + II) | 4 | 99 | 5 | 146 |
| RESP0381 | m | 8 | 1a_aed_eq | E | Low Grade Tumor (WHO I + II) | 6 | 13 | 10 | 19 |
| RESP0384 | f | 13 | 1a_aed_stop | E | cavernoma | 16 | 95 | 3 | 13 |
| RESP0396 | f | 15 | 1a_aed_low | E | Low Grade Tumor (WHO I + II) | 9 | 44 | 5 | 73 |
| RESP0409 | f | 12 | 1a_aed_stop | E | FCD | 5 | 9 | 2 | 20 |
| RESP0428 | m | 19 | 1a_aed_low | E | Low Grade Tumor (WHO I + II) | 16 | 53 | 18 | 99 |
| RESP0451 | m | 19 | 1a_aed_stop | E | FCD | 17 | 29 | 39 | 18 |
| RESP0465 | f | 11 | 1a_aed_eq | E | gliosis/scar | 5 | 13 | 6 | 109 |
| RESP0586 | f | 28 | 1a_aed_eq | E | cavernoma | 6 | 24 | 1 | 22 |
| RESP0619 | m | 14 | 1a_aed_stop | E | gliosis/scar | 9 | 14 | 8 | 27 |
| RESP0634 | m | 19 | 1a_aed_stop | E | gliosis/scar | 9 | 43 | 8 | 39 |
| RESP0059 | f | 25 | 1a_aed_eq | T | MTS | 9 | 25 | 29 | 20 |
| RESP0087 | f | 51 | 1a_aed_eq | T | cavernoma | 11 | 93 | 24 | 78 |
| RESP0094 | m | 72 | 1a_aed_eq | T | malformation cort. development | 13 | 34 | 51 | 27 |
| RESP0096 | m | 25 | 1a_aed_eq | T | MTS | 0 | 16 | 2 | 72 |
| RESP0105 | f | 13 | 1a_aed_low | T | Low Grade Tumor (WHO I + II) | 14 | 24 | 32 | 21 |
| RESP0150 | m | 44 | 1a_aed_eq | T | no abnormalities | 6 | 46 | 19 | 34 |
| RESP0269 | f | 13 | 1a_aed_eq | T | gliosis/scar | 5 | 35 | 31 | 36 |
| RESP0280 | m | 13 | 1a_aed_low | T | Low Grade Tumor (WHO I + II) | 6 | 25 | 40 | 36 |
| RESP0282 | f | 20 | 1a_aed_low | T | gliosis/scar | 4 | 34 | 31 | 55 |
| RESP0283 | m | 31 | 1a_aed_low | T | Low Grade Tumor (WHO I + II) | 0 | 64 | 34 | 32 |
| RESP0284 | m | 19 | 1a_aed_low | T | Low Grade Tumor (WHO I + II) | 5 | 37 | 28 | 31 |
| RESP0316 | m | 19 | 1a_aed_low | T | MTS | 6 | 72 | 21 | 46 |
| RESP0332 | f | 17 | 1a_aed_stop | T | Low Grade Tumor (WHO I + II) | 5 | 27 | 36 | 23 |
| RESP0349 | m | 24 | 1a_aed_eq | T | cavernoma | 7 | 33 | 23 | 31 |
| RESP0356 | m | 22 | 1a_aed_eq | T | TuberoSclerosis | 5 | 31 | 24 | 24 |
| RESP0433 | m | 22 | 1a_aed_stop | T | Low Grade Tumor (WHO I + II) | 6 | 22 | 8 | 23 |
| RESP0437 | m | 16 | 1a_aed_stop | T | Low Grade Tumor (WHO I + II) | 6 | 6 | 15 | 21 |
| RESP0462 | f | 24 | 1a_aed_low | T | MTS | 14 | 35 | 44 | 26 |
| RESP0464 | f | 21 | 1a_aed_stop | T | Low Grade Tumor (WHO I + II) | 4 | 25 | 28 | 29 |
| RESP0475 | f | 15 | 1a_aed_stop | T | MTS | 6 | 38 | 18 | 28 |
| RESP0546 | f | 19 | 1a_aed_stop | T | Low Grade Tumor (WHO I + II) | 10 | 33 | 30 | 27 |
| RESP0557 | m | 28 | 1a_aed_eq | T | cavernoma | 5 | 0 | 94 | 38 |
| RESP0570 | f | 64 | 1a_aed_eq | T | FCD | 8 | 32 | 24 | 29 |
| RESP0578 | m | 59 | 1a_aed_low | T | FCD | 7 | 20 | 38 | 5 |
| RESP0579 | m | 24 | 1a_aed_eq | T | gliosis/scar | 14 | 46 | 30 | 20 |
| RESP0595 | f | 36 | 1a_aed_low | T | cavernoma | 2 | 0 | 32 | 27 |
| RESP0599 | f | 61 | 1a_aed_eq | T | gliosis/scar | 5 | 26 | 36 | 31 |
| RESP0625 | f | 9 | 1a_aed_stop | T | Low Grade Tumor (WHO I + II) | 5 | 21 | 45 | 23 |
| RESP0645 | f | 5 | 1a_aed_low | T | Low Grade Tumor (WHO I + II) | 3 | 15 | 15 | 31 |
| RESP0673 | f | 62 | 1a_aed_low | T | cavernoma | 8 | 29 | 25 | 28 |

### Measuring effect across all the channels

Figure 1 shows the comparison between the biomarker distributions of values computed in pre-resection resected channels in improved patients (Engel 1A) and post-resection channels in cured patients (Engel 1A without medication). Five out of seven biomarkers (ARR, PAC,PLI, H2, GC) were significant (p < 0.01) using a one-sided Kolmogorov-Smirnov test.

**Figure 1.**
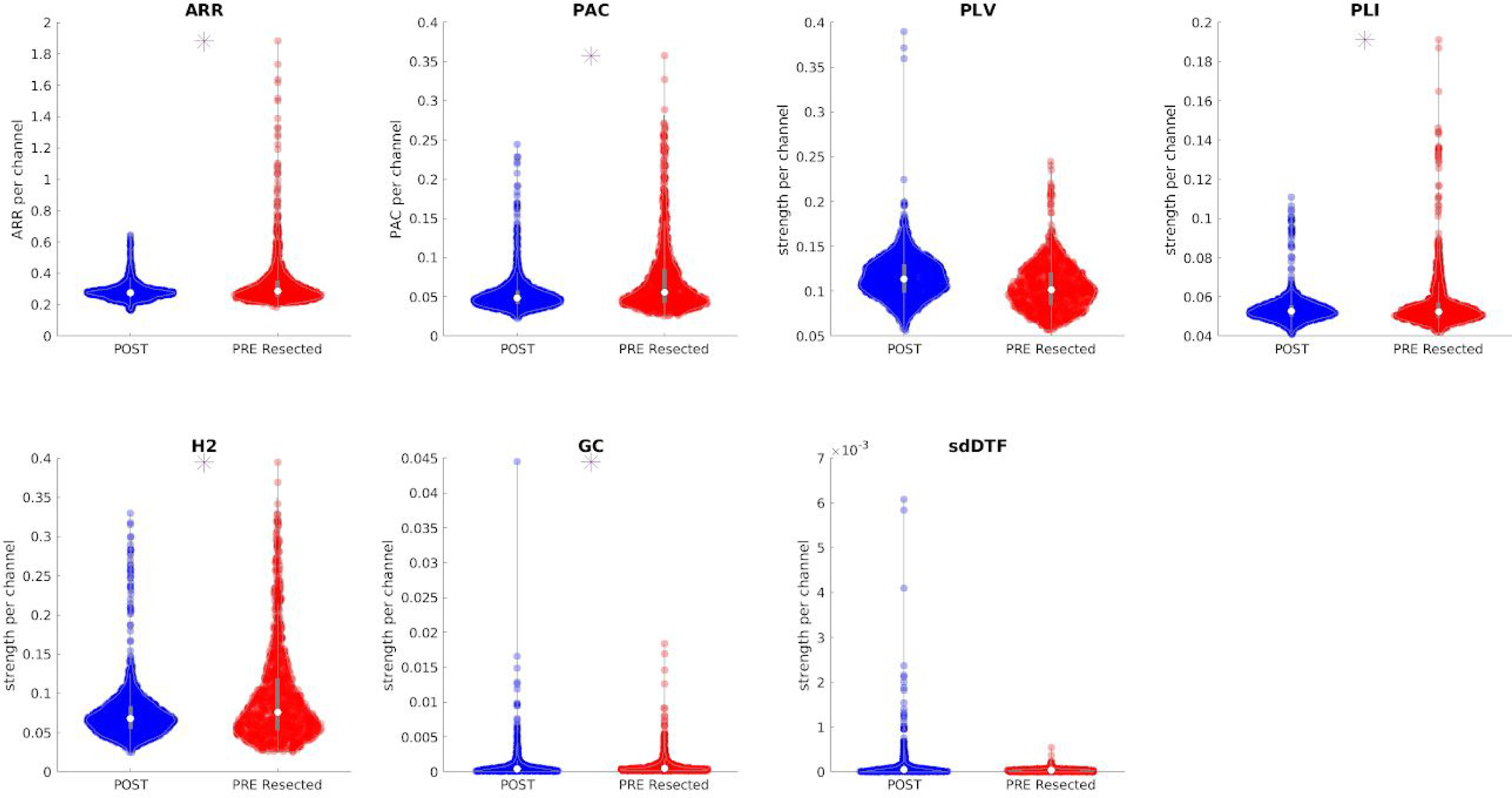
Comparison between biomarker distributions values computed on pre-resection resected channels (red) and post-resection channels (blue) in cured patients. The presence of an asterisk indicates that the two distributions are significantly different (p < 0.01 one-sided Kolmogorov-Smirnov test). Note that each point for the univariate biomarkers (ARR and PAC) represents the value of the biomarker per channel, while the y-axis for the bi-/multi-variate biomarkers represents the strength. Inside each violin-plot a boxplot is depicted in gray with the median value highlighted with a white dot.

### Measuring effect using maximum per patient

Figure 2 shows the comparison between the distribution of maximum values computed in pre-resection resected channels for improved patients and maximum post-resection channel values for cured patients. We show the five biomarkers for which a significant effect was reported across all the channels (Figure 1). Each coloured dot represents the maximum value of the biomarker for each individual patient across all the channels and situations. Although for all the biomarkers the pre-resection resected distribution has a longer tail than the post-resection one, only the distributions related to PAC are significantly different (p < 0.01).

**Figure 2.**
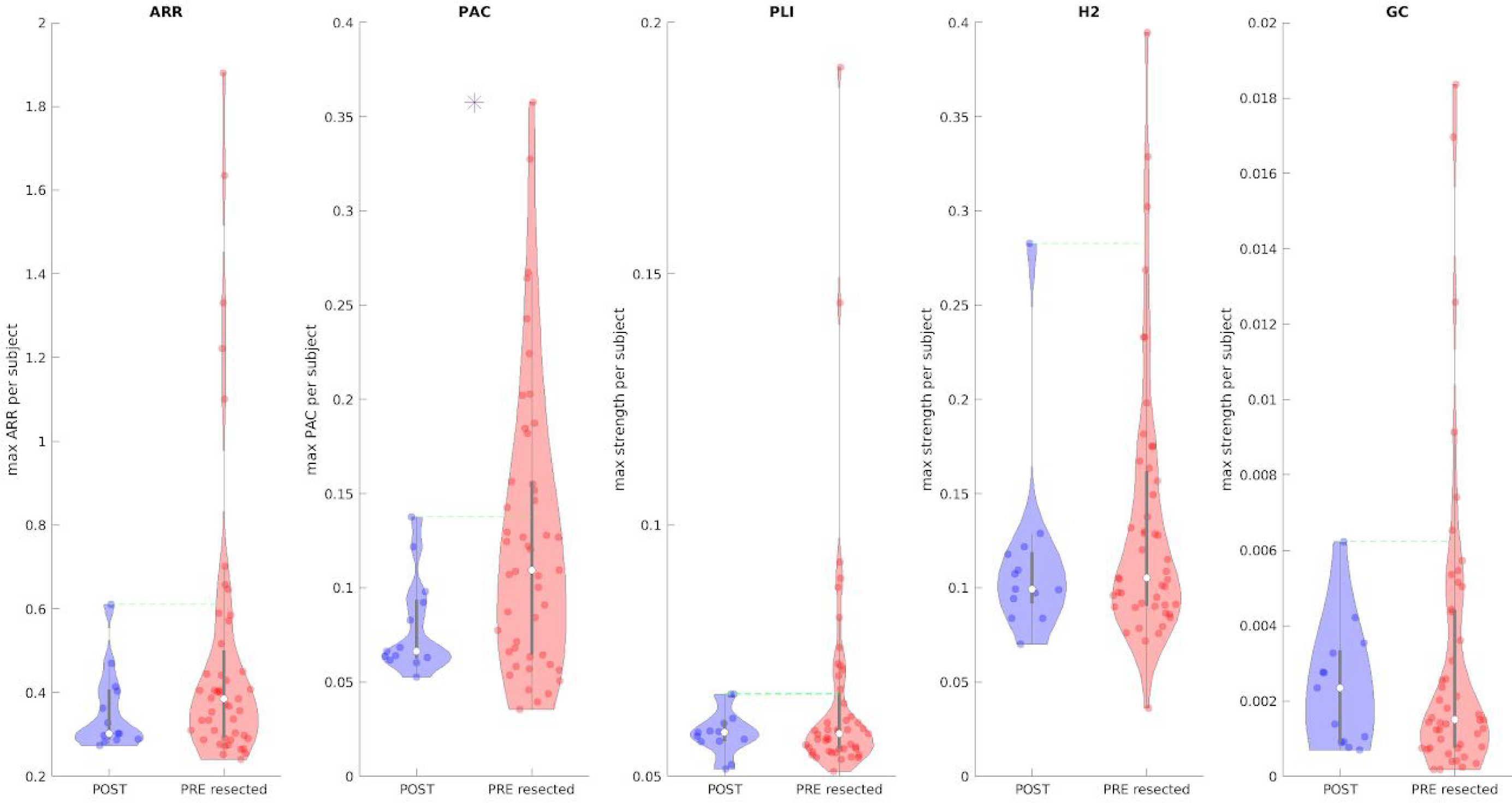
Comparison between maximum biomarker values between pre-resection resected channels (red) in improved patients and post-resection channels in cured patients (blue). Each dot represents the maximum value of the biomarker across all channels of each patient. The presence of an asterisk indicates that the two distributions are significantly different (p < 0.01 one-sided Kolmogorov-Smirnov test). Inside each violin-plot a boxplot is depicted in gray with the median value highlighted with a white dot. For each biomarker, the green line represents the threshold used to define the normal tissue (biomarker reference) using post-resection cured patients.

We defined a threshold for each biomarker as the maximum across all patients in the post-resection distribution so that we could quantify the number of patients for whom in the pre-resection recordings we could localize the channel to be resected. The best performance was obtained using PAC, 16 out of 47 patients are above the threshold (a sensitivity of 34%).

### Pooling together all the biomarkers

Figure 3 shows for each patient in the pre-resection recordings the number of biomarkers above their respective thresholds (i.e. the maximum across all channels and all patients in the post-resection cured group). For each patient, we counted if at least one channel showed a value higher than the respective threshold for each biomarker. The biomarker counts are higher in temporal patients compared to extra-temporal ones. There was no patient with all the biomarkers above the thresholds. These two results suggested that the biomarkers convey different kind of information related to epileptogenicity.

**Figure 3.**
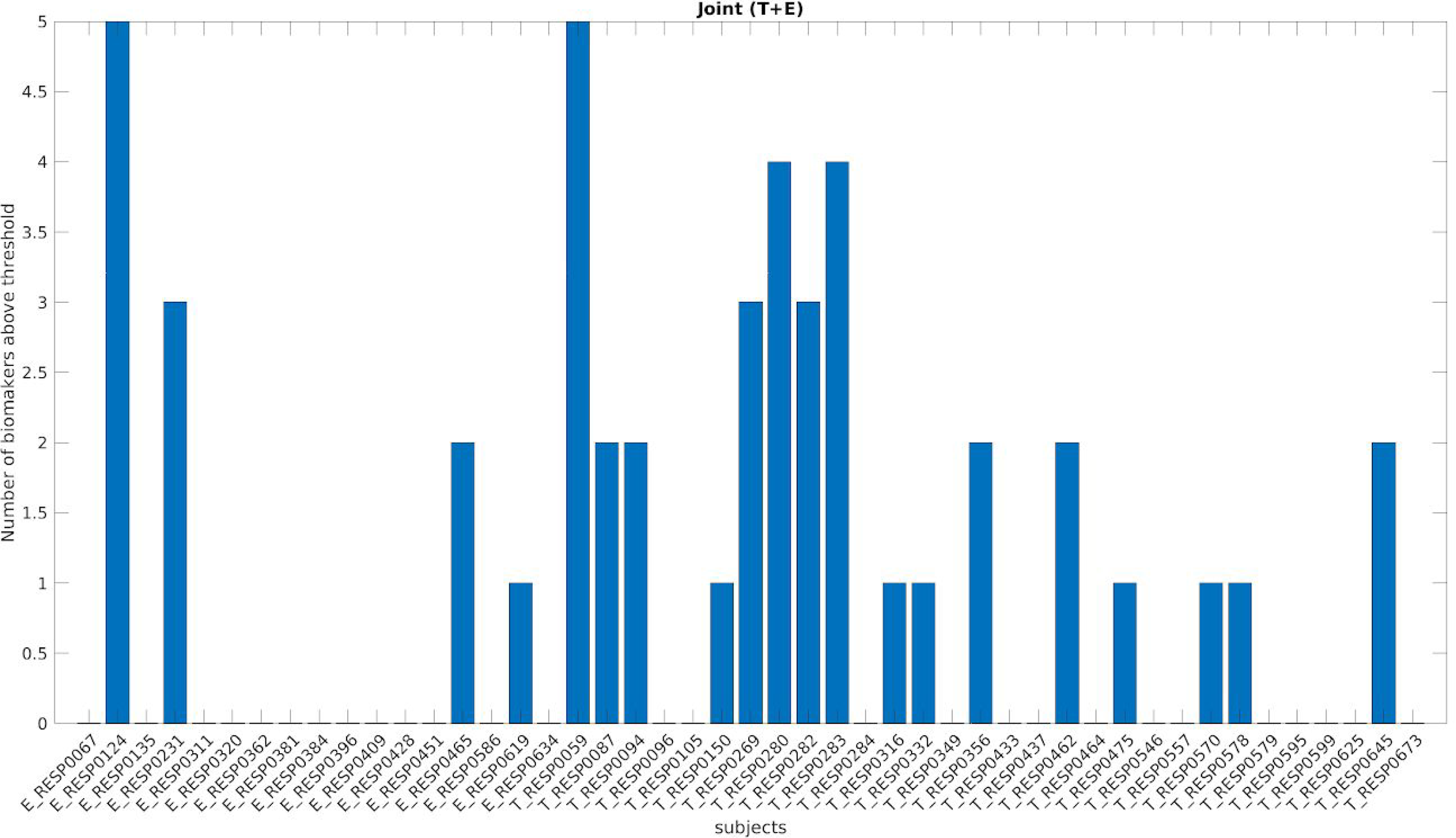
Number of biomarkers above the threshold for each patient. On the x-axis each of the 47 improved patients is displayed with a coded number. The first 17 patients are extra-temporal patients (E before the coded name), while the remaining 30 patients are temporal patients (T before the coded name). The y-axis represents the number of biomarkers above the specific threshold (computed separately for each biomarker) for each patient.

If we defined a new biomarker (‘cumulative’ biomarker) combining together the contribution of all biomarkers (i.e. at least one biomarker above the threshold), we obtained a performance of 20 out of 47 patients (sensitivity of 42%) improving of 8% the best sensitivity found using one single biomarker.

We decided to investigate separately the two subgroups of temporal and extra-temporal patients, recomputing all the threshold for each biomarker independently for each subgroup.

Figure 4 and Figure 5 show the number of biomarkers above the thresholds for each patient in the temporal and extra-temporal subgroup. After recomputing the threshold independently for each subgroup, it seems that the biomarkers are more sensitive in the temporal subgroup. Figure 6 summarizes the performances of the ‘cumulative’ biomarker for the whole group, the temporal and extra-temporal subgroups. The best performance was obtained considering the temporal subgroup: 28/30 (sensitivity of 93%) patients showed a value in the pre-resection recordings higher than the threshold for at least one biomarker of the pool of biomarkers. However, for the extra-temporal patients only 4 out of 17 patients (sensitivity of 23%) were above the threshold.

**Figure 4.**
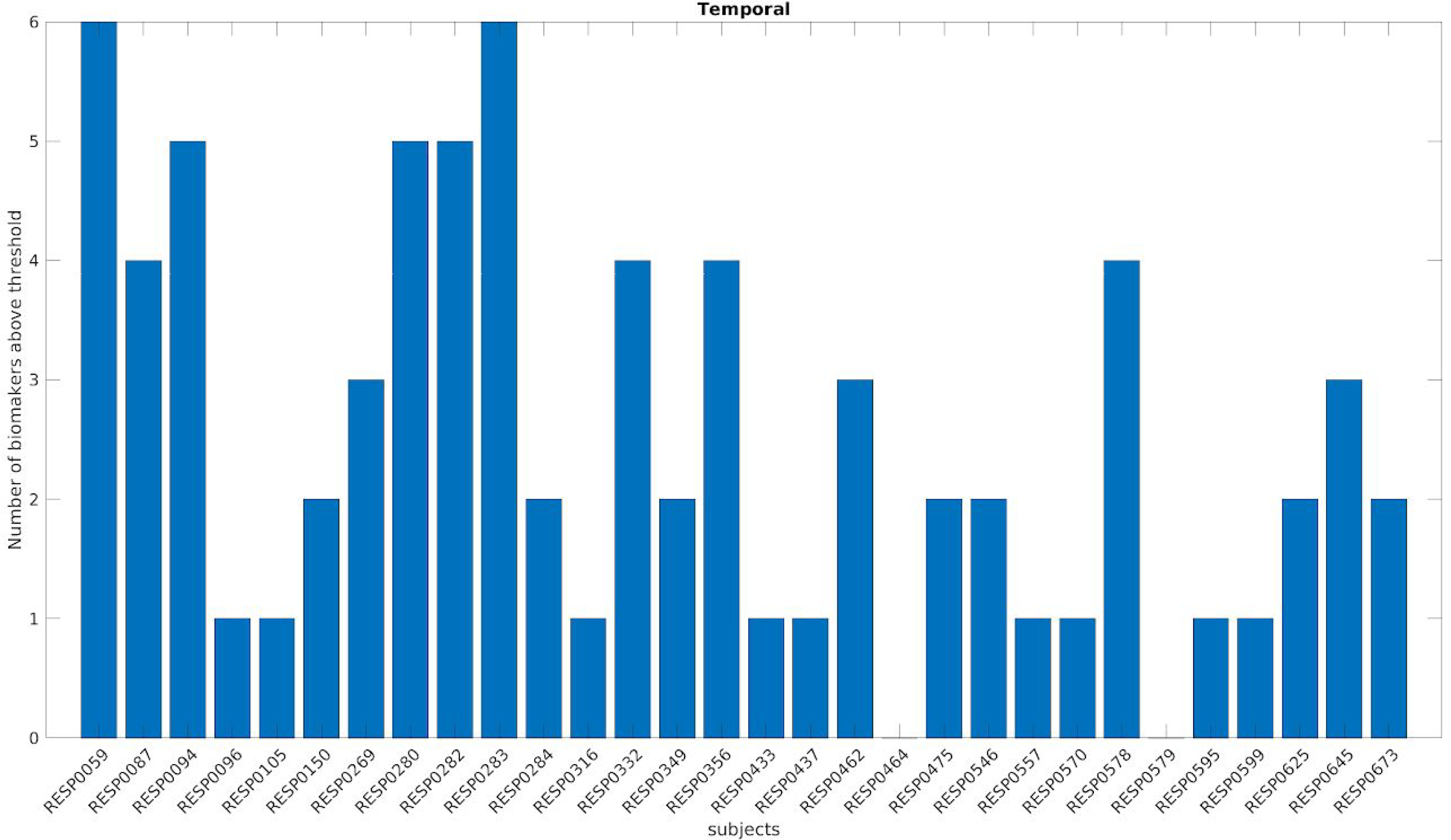
Number of biomarkers above the threshold for each temporal patient. On the x-axis each of the 30 improved temporal patients is displayed with a coded number. The y-axis represents the number of biomarkers above the specific threshold for each patient (computed independently for each biomarker and using only the temporal patients).

**Figure 5.**
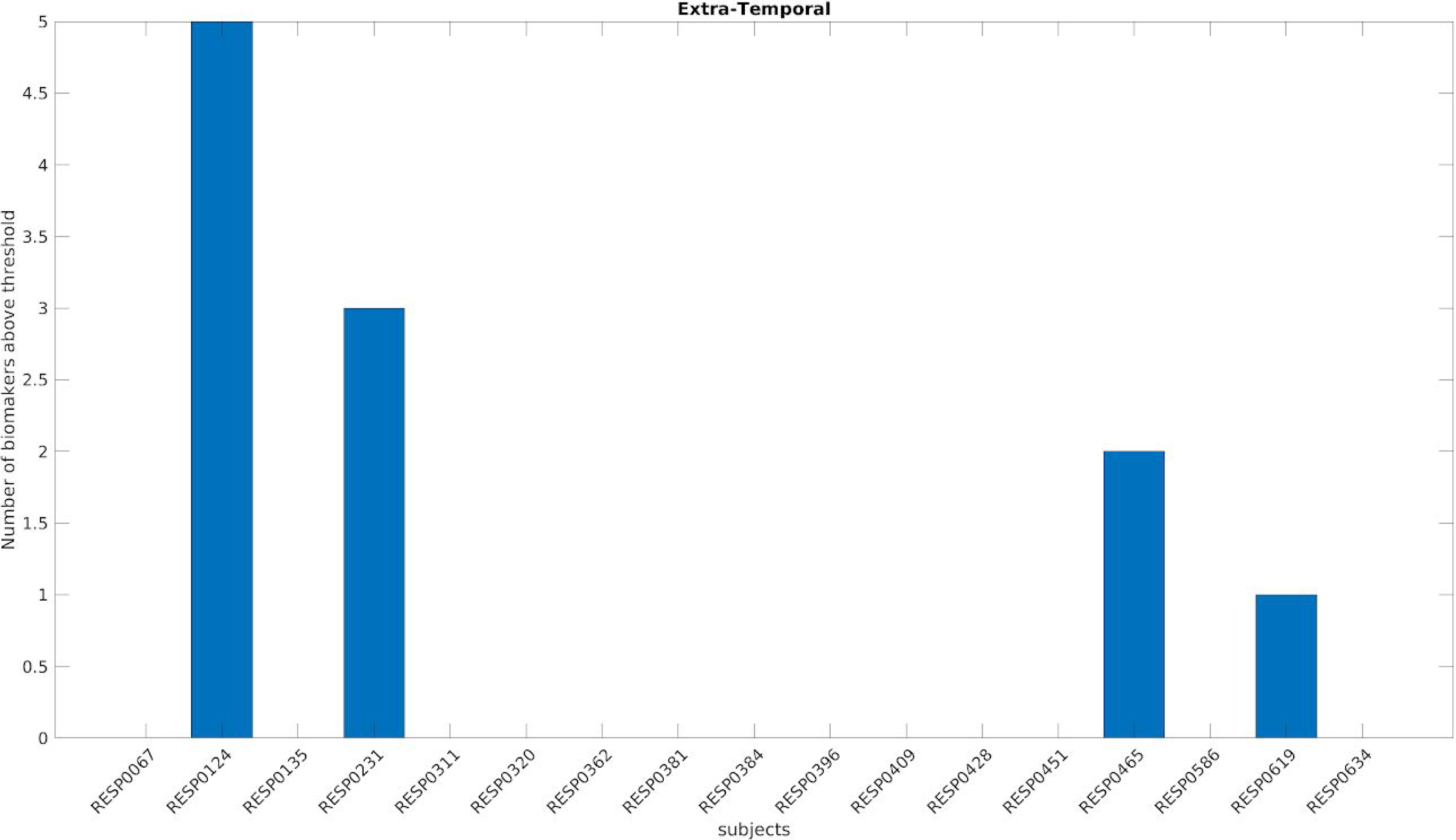
Number of biomarkers above the threshold for each extra-temporal patient. On the x-axis each of the 17 improved extra-temporal patients is displayed with a coded number. The y-axis represents the number of biomarkers above the specific threshold for each patient (computed independently for each biomarker and using only the extra-temporal patients).

**Figure 6.**
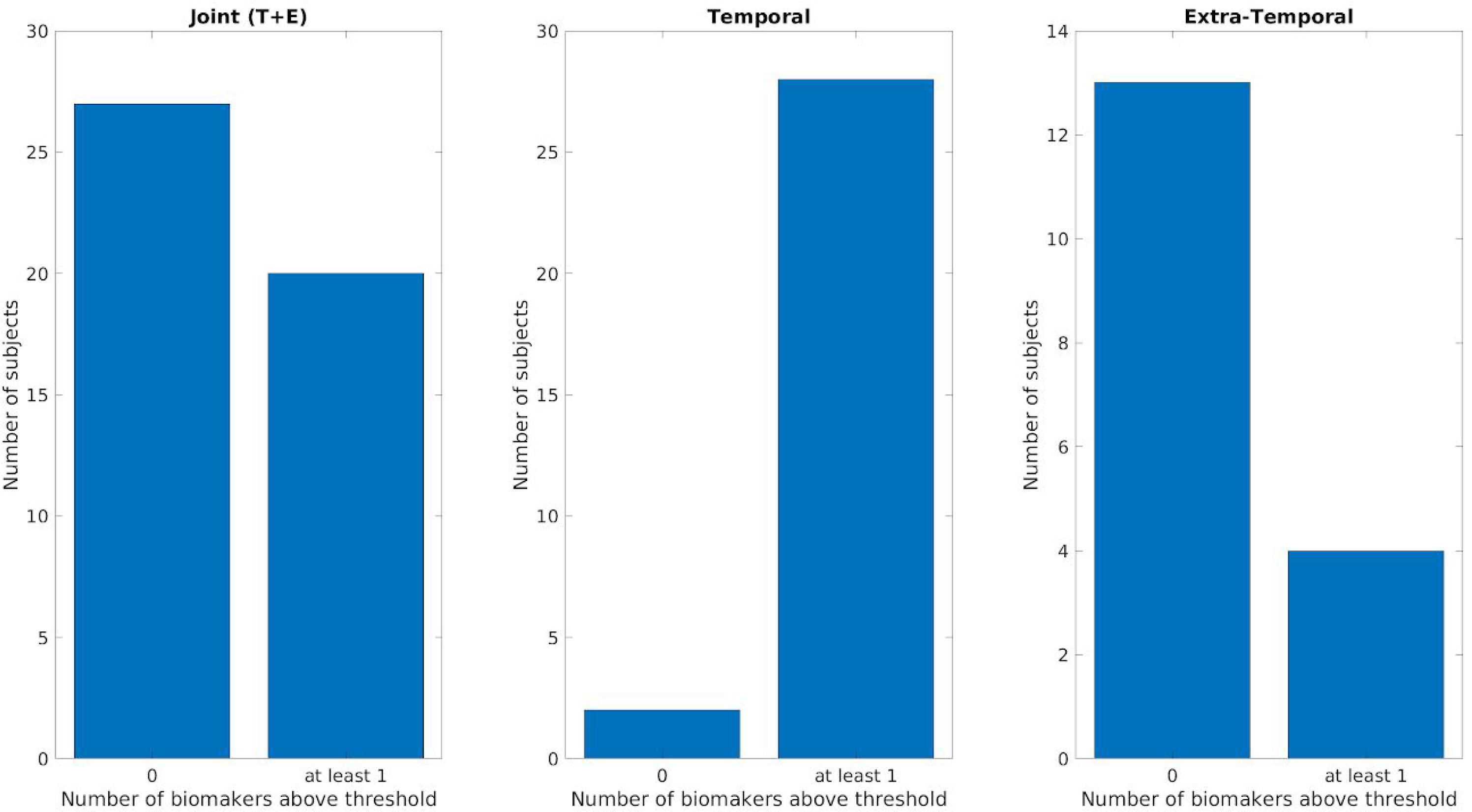
Comparison of the ‘cumulative’ biomarker for the three different groups (Joint Temporal and Extra-Temporal, only Temporal, only Extra-Temporal). Y-axis counts the number of patients considering the pre-resection recording of resected channels for which none (failed detection, labeled as 0 on x-axis) or at-least one biomarker (detection, labeled as at least 1) is above its respective threshold.

### Mesiotemporal versus neocortical channels

In temporal patients we found a significant (p<0.01) difference between mesiotemporal channels and neo-cortical channels for the PAC and GC, while no significant difference was found for the remaining biomarkers.

## Discussion

This study investigates the performances of different univariate, bivariate and multivariate signal biomarkers, used separately and combined, to discriminate between healthy and epileptogenic tissue using inter-ictal data derived from ioECoG. We performed all the analyses in a ground-truth scenario, using post-resection recordings of completely cured patients (for whom seizure control without medication was achieved for at least one year after resection) as a way to define a reference threshold for healthy tissue to be compared with channels in pre-resection recordings of improved patients.

We chose our biomarkers with two criteria in mind: 1) to be exhaustive regarding the different types of measures used (i.e. univariate, bivariate and multivariate); 2) biomarkers should have been reported to show an overall significant effect in discriminating between healthy and pathological tissue using inter-ictal intracranial recordings^23–25,27–29,31–35,60^.

We could replicate previous findings regarding the detection of an overall effect, normal versus abnormal tissue, comparing separately the distribution of pre-resection recordings against post-resection recordings in 5 out of 7 biomarkers. This is a remarkable result considering the differences in methodological (and arbitrary) choices we used to harmonize the analysis pipelines of the aforementioned studies. For the biomarkers for which we failed to observe a significant effect (PLV and sdDTF) this failure could be indeed related to different signal processing pipelines regarding the epoch length^61^, the reference montage^62,63^, the different state of vigilance^64–66^, which could have affected the results.

The resection area in good seizure outcome patients often includes normal brain tissue along with electrophysiologically abnormal tissue. In order to overcome this problem, we repeated our analysis using the maximum value across channels for each subject. Although each tested biomarker showed a longer tail of the pre-resection values compared to the post-resection values, only the PAC could detect a significant difference between the two distributions. Furthermore, it revealed the best sensitivity allowing to detect the pathological tissue in 16 out of 47 patients. Our results confirm the important role of cross-frequency coupling in neuronal communications^43,67^ and also reinforce the idea that abnormal PAC values are linked to ictogenesis^32–35,60,68–71^.

A possible explanation of the failure of the other biomarkers might be the fact that the maximum across channels represents a too strict and crude statistic to detect an effect. In fact, the maximum statistic works on the implicit assumption that one channel with an electrophysiologically abnormal value (i.e. higher value than the threshold) is enough for ictogenesis. However, evidence has been accumulated on the role of a critical mass to trigger seizures, hence the maximum as a statistic may overlook important global network features that go beyond single channel statistics^72–77^.

The combination of the whole pool of biomarkers improved the sensitivity (from 34% to 42%) in terms of patients for whom it was possible to detect pathological tissue in the pre-resection recordings: 20 out of 47. This result suggests that different biomarkers may capture different mechanism of ictogenesis and it is inline with recent literature suggesting that more robust results are shown by combining different biomarkers^33,34,78^ since they potentially exploit independent information.

When we performed separately the analysis depending on the type of epilepsy (temporal or extra-temporal), the combination of multiple biomarkers for temporal patients held the remarkable result of 93% sensitivity, while 23% was obtained for the extra-temporal group. This performance difference may point out different structure related mechanisms (i.e. neo-cortical versus mesiotemporal) involved into the ictogenesis to which biomarkers may be responsive or not. Hence, considering separately the two groups allows for a definition of better threshold (i.e. more structure tuned) to discriminate between normal and pathologic tissue. However, it could be that this result is influenced by the different neurophysiological properties of the tissue independently from the epileptogenicity. The biomarkers are detecting a difference in terms structure (mesiotemporal vs neocortical) rather than epileptogenicity. A recent published intracranial ECoG atlas^79^ of recording in healthy tissue points into this direction, highlighting how different anatomical brain areas have specific electrophysiological signatures in terms of spectral oscillatory and non-oscillatory properties. Indeed, we found a significant difference (PAC and GC) in temporal patients comparing mesiotemporal channels with neocortical ones. However, the sensitivity of 93% cannot be fully explained in structure related terms since for temporal patients the maximum value above the biomarker reference was found in the mesiotemporal channels 11 times out of 18 for PAC and 5 out of 13 for GC.

Nevertheless, our results regarding temporal patients are comparable to a recent similar work on epileptogenic localization in which good performances are obtained on a dataset of predominantly temporal patients^34^. The poor performances in the extra-temporal group could also be affected by the limited amount of resected channels considered in the pre-resection recordings. The mean number of resected channels available in the extra-temporal group was around 13 compared to 30 in the temporal group, and for two extra-temporal patients only one channel was available.

It is important to realize that, in this retrospective study, the total amount of data analyzed per situation (1 minute) represents a drawback since it has been shown that longer periods of data are needed to detect pathological signatures^33,34,65^. However, the time constraint in intra-operative recordings will be always an issue, because the goal is not only to find a biomarker able to discriminate what is normal/abnormal but it is to accomplish it in a reasonable amount of time relative to surgery duration.

The choice of solely the gamma frequency band for some of the biomarkers is a limitation, since it has been reported that brain networks findings in intracranial recordings are frequency-dependent^22,80^. However, our a priori choice was motivated by previous works in which gamma frequency band appeared consistently to reveal significant results using inter-ictal intracranial recordings^22,23,26,28^.

In this study there are three main limitations related to localization matters. The first, consists of the not straightforward way to project the bivariate and multivariate biomarkers computed from signals recorded from two (or more) different locations to a single location. This is not an issue for univariate measures since they provide a more confined measure, in terms of localization. However, the employment of a bipolar montage, even though to a lesser extent, posit the same obstacle. The use of high density grids can be a possible approach to improve the localization precision.

The second, is related to the unavailability of accurate electrode localization to compare the value of the biomarkers on the same tissue pre- and post-resection (what is left after resection). In fact, using the pre- and post-resection pictures is enough to mark (not-)resected channels, but it does not allow to quantify the value of the biomarker in the same location pre- and post-resection. Third, in temporal patients, there is uncertainty on the part of the mesiotemporal structure we are recording from since we do not have the exact position of the placement the strip.

In conclusion, in this retrospective study, using a substantial number of patients for whom seizure control was improved one year after the operation, we pointed out the importance to work on a ground-truth scenario to evaluate biomarker performance in an unbiased way. Our results suggested that a universal unique biomarker is insufficient to pinpoint the epileptogenic tissue. The combination of different biomarkers improved the localization performances, however the results should be considered more from a perspective of pathophysiological understanding rather than as a tool for the operation theater since sensitivity achieved is not adequate.

## Data Availability

no sharing of the data

## Appendix

### Auto-Regressive Residual Modulation

Given *x* as a signal we divided the signal in overlapping windows *w*_1_ … *w*_*n*_. For each window *w*_*i*_ it is possible to compute the auto-regressive model of order (in Geertsema’s work^30^ the model order was 3) for every time point with the following formula:

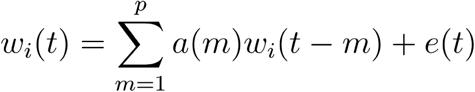

where *a*(*m*) are the coefficient of the model, *e*(*t*) is the residual for each time point *t*. Then, it is possible to compute the variation of the residual per window *w*_*i*_ and order *p* as 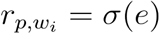. The auto-regressive residual modulation can be computed as the coefficient of variation (CV) of the residuals across the windows:

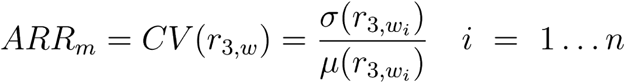

where 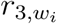 is the residual variation for window *w*_*i*_ for model order 3. Geertsema et al. ^31^ suggested an improved version of *ARR*_*m*_. This modified version has been shown to be less sensitive to artefacts compared to the original version^30^. The authors noticed that the decline of the residuals of different model orders (order 1 and 2) was different for artefacts compared to real events (spikes, high frequency oscillations). Therefore, they use the steepness of the residual decline from the first order 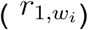 to the second order 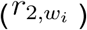 to filter the residuals with order 3 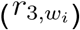 to include in the computation of the CV. Specifically, they computed the residual decline *D*(*w*_*i*_) for each window *w*_*i*_ as:

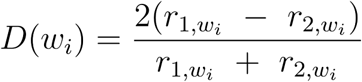

Then, they defined the residual of model 3 for a specific window an outlier 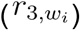 if the following two criteria were satisfied:

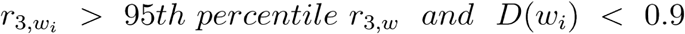

In that case the residual variations of window and contiguous windows were removed 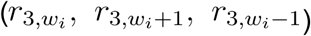. This provides selection of cleaned windows over which compute the modified version of ARRm:

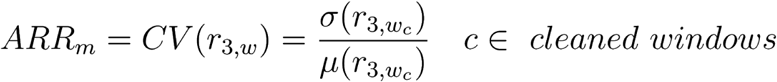

### Phase Amplitude Coupling

Given *x* as a signal, we filtered the signal in two frequency bands, theta band (4-8Hz) and gamma band (30-80Hz) obtaining two filtered signals *x*_*θ*_ and *x*_*γ*_. We then computed the Hilbert transform of the two filtered signals to obtain the instantaneous phases and amplitude envelopes for each time point *t, ϕ*_*θ*_(*t*), *A*_*θ*_(*t*), *ϕ*_*γ*_(*t*), *A*_*γ*_(*t*). We then compute PAC with the following formula:

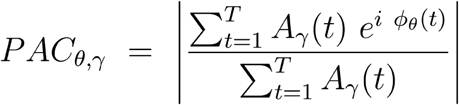

Where *T* is the signal length in samples, is the absolute operator.

### Phase Locking Value

Given *x* and *y* as two signals, we filtered the signals in gamma frequency band (30-80Hz), obtaining two filtered signals *x*_*γ*_ and *y*_*γ*_. We then computed the Hilbert transform of the two filtered signals to obtain the instantaneous phases *ϕ*_*x,γ*_(*t*), and *ϕ*_*y,γ*_(*t*) and for each time point *t*. We then compute PLV with the following formula:

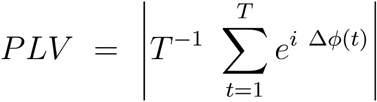

where Δ*ϕ*(*t*) = *ϕ*_*x,γ*_(*t*) - *ϕ*_*y,γ*_(*t*), *T* is the signal length in samples, | | is the absolute operator.

### Phase Lag Index

Given *x* and *y* as two signals, we filtered the signals in gamma frequency band (30-80Hz), obtaining two filtered signals *x*_*γ*_ and *y*_*γ*_. We then computed the Hilbert transform of the two filtered signals to obtain the instantaneous phases *ϕ*_*x,γ*_(*t*), and *ϕ*_*y,γ*_(*t*) for each time point *t*. We then compute PLI with the following formula:

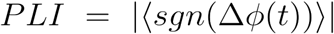

where *Δϕ*(*t*) = *ϕ*_*x,γ*_(*t*) - *ϕ*_*y,γ*_(*t*), < > is the average across all time points *t*, | | is the absolute operator and *sng* is the sign function:

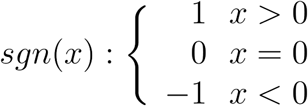

### Non linear correlation coefficient

The implementation of *h*^2^ used in this work follows the implementation suggested by Kalitzin et al.^81^ that is obtained using the following formula:

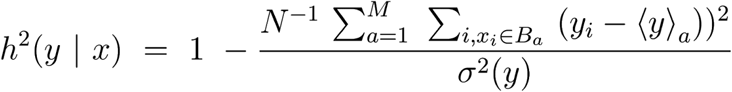

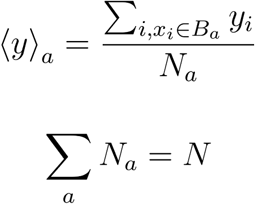

Where *N* is the time length of the two signal *x*_*i*_ and *y*_*i*_ for *i* = 1 …*N*. The values of *x*_*i*_ are binned in *M* bins *B*_*a*_ with *a* = 1 …*M* each containing *N*_*a*_ points. *h*^2^(*y* | *x*) represents the variation of *y* explained by *x*, while *h*^2^(*x* | *y*) the variation of *x* explained by *y*.

### Granger Causality

Granger causality index for a bivariate situation in which there are two time-series *x* and *y* can be defined as the logarithm of the ratio between the autoregressive residual considering the model with one variable *x* over the autoregressive residual value of the model with two variable *x* and *y*:

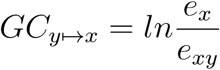

where *e*_*x*_ and *e*_*xy*_ are the residual variance from the autoregressive model using only previous values of and the residual variance using previous values of *x* and *y*.

This definition can be generalized to a multivariate (multi-channels) case with the following formula:

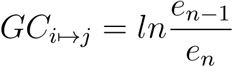

where Granger causality from *i* to *j* is equal to the natural logarithm of the ratio between the variance of the residual using the reduced regressive model (considering all the time-series *n* - 1 other than *i*) and the variance of the residual obtained from the full model (considering all the *n* time series). For the time based GC we used the implementation in the MVGC toolbox^59^.

### short-time direct Directed Transfer Function

The sdDFT is can be computed using the following formula^82^:

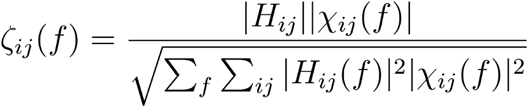

where *H*_*ij*_ is the transfer function describing the directed causal relationship from *j* to *i* at frequency *f, χ*_*ij*_ is the direct partial coherence between *i* and *j*. The combination between the *H*_*ij*_ and *χ*_*ij*_ gives a measure of directed causal interaction between *j* and *i* in a multivariate (multi-channel) system.

We used the implementation in the SIFT toolbox for sdDTF^57,58^.

## Acknowledgments

M. Demuru was supported by the TKI Health Holland grant LSHM16054-SGF.

M. Zijlmans was supported by the ERC starting grant 803880

